# Use and Perceptions of AI Chatbots for Mental Health Support Among Adults with Lived Experience

**DOI:** 10.64898/2026.07.11.26357785

**Authors:** Haruka Notsu, Phuong Anh Nguyen, Matthew Flathers, Sean Ryan, Jill Noorily, Leah Wentworth, Christine Crawford, Michael Wood, Daniel Gillison, John Torous

## Abstract

**Importance:** AI chatbots are increasingly used for mental health support, but little is known about how adults with lived experience of mental health condition use and perceive these tools.

**Objective:** To characterize the use and perception of AI chatbots, including for mental health purposes, among adults connected to a large US mental health organization,

**Design:** Cross-sectional online survey conducted from March to May 2026.

**Setting:** Adults recruited through email newsletters from the National Alliance on Mental Illness (NAMI), the largest grassroots mental health organization in the US.

**Participants:** Adults aged 18 years older with English proficiency. Affiliation with NAMI or a diagnosis of mental health disorder was not required.

**Results:** Of 454 participants, 316 (69.6%) reported having used an AI chatbot. Use was more common among younger participants and those with a current mental health diagnosis. Among AI users, 133 (42.1%) reported using a chatbot for mental health purposes. Mental health-related use was typically brief and focused on information gathering and in-the-moment emotion regulation. Most users rated chatbots as helpful for their mental health. Among the 95 participants with a mental health provider, only 14 (14.7%) had openly discussed their AI use with their provider. Higher frequency of AI use was associated with greater odds of disclosure (OR, 1.67; 95% CI, 1.20-2.38; *P* = .003).

**Conclusion and Relevance:** In this survey of adults connected to a large mental health organization, AI chatbots were widely used but engagement for mental health purposes was typically brief and focused. Most use occurred without clinician awareness, suggesting a need for proactive conversations about AI use in routine mental health care.

**Key Points:** *Question:* How do adults with lived experience of mental health condition use and perceive AI chatbots for mental health purposes?

*Findings:* In this cross-sectional survey of 454 adults, 133 reported using one for mental health purpose. Use was more common among participants with a current diagnosis. Most use was brief and focused. Only 14.7% of those with a mental health provider had openly discussed their AI use.

*Meaning:* Among adults with lived experience, AI chatbots may function as an accessible tool for information and immediate coping rather than a replacement of therapy, but this use is largely undisclosed to clinicians.

---

A paradigm shift in the delivery of mental health services, fueled by Artificial Intelligence (AI) chatbots, is underway. The growth of AI in healthcare and mental health is impossible to ignore. A recent national poll reports 32% of US adults reported using AI chatbot for health information in the past year, with 16% specifically for mental health [1]. Another national survey reports that more than half of US teens using AI chatbots to search for information, with 12% using them for emotional support [2]. But while the broader trend is clear, less is known about how people with mental health conditions themselves perceive and use AI chatbots.

The need for a more focused understanding of real-world AI use cases for mental health is clear. While there has been some industry led research examining how people may use mental-health-specific AI product [3], the vast majority of people use general-purpose AI models (e.g. ChatGPT, Gemini, Claude) and these models may already be acting as the single largest provider of mental health support today [4]. But we do not know how people with mental health conditions engage with these tools (e.g. for psychoeducation vs psychotherapy), whether they find them harmful or helpful, and even if rates of use parallel general population estimates. With interest from regulators, clinical teams, and even the technology vendors themselves to make AI for mental health safer [5], now is the time to produce the real-world data that can be used to guide better and safer AI tools. Such research should be led by the voice of lived experience, specifically individuals with mental health conditions, their family members and caregivers. To provide a more rigorous empirical foundation for understanding how AI chatbots are being adopted for mental health support, we conducted a cross-sectional survey of adults recruited through our partnership with the National Alliance on Mental Illness (NAMI), the largest grassroots mental health organization in the US NAMI offers education, advocacy and support through over 650 State Offices and local affiliates nationwide, and provides a HelpLine for individuals with mental health concerns and family members seeking information and support about mental health. NAMI’s programming is available at no cost to the participant, and NAMI serves millions of individuals through the work of the Alliance and the NAMI National office. While NAMI classically does not conduct research outside of evaluating their own programs and initiatives, the urgency of assessing real-world AI uses prompted this collaborative project.

## Methods

### Participants

Participants were adults recruited through NAMI email lists for volunteers, staff, and other individuals committed to supporting mental health. This included individuals with lived experience of mental illness, mental health and public health professionals, family members and friends, and community members who are dedicated to improving the lives of those affected by mental illness.

Participation was voluntary, with no compensation. Eligibility criteria were: 18 years of age or older, English proficiency, and ability to understand and provide informed consent.

### Survey Design

The survey instrument was developed by the study team, with additional rounds of edits from people with lived experience, and was built on the structure of a prior self-report survey on AI and mental health [6]. The final instrument consisted of 55 items organized into sections: (1) demographic questions; (2) general AI chatbot usage (e.g., tools used, frequency, duration); (3) mental-health-specific AI chatbot usage (e.g., specific activities, frequency, perceived trustworthiness and help/harm); (4) non-user items (e.g., reasons for non-use, perceived best uses). Branching logic was used to tailor follow-up questions based on participants’ responses to questions. General and mental-health-specific usage questions were only displayed for participants who indicated using AI chatbots for these purposes. Non-user items are displayed only to participants who indicated they never use AI for any purpose. Items used single-select, multi-select, and free-text response formats. The median survey completion time for this sample was 3.5 minutes (interquartile range: 2.4 -5.0). A full copy of the survey is accessible in Appendix 1.

### Study Procedures

A description of the study was included in two newsletters, one for a general NAMI audience, and one for youth, young adults, and individuals interested in youth mental health. An email reminder about the study was also distributed to the mailing lists with a brief description of the study and a link to the online survey. The survey was administered online through REDCap, a secure web-based survey platform from March to May, 2026. The first page of the survey contained the informed consent page. All procedures were approved by the Beth Israel Deaconess Medical Center Institutional Review Board (2026P000064).

### Data Analyses

Analyses were primarily descriptive, following the Strengthening the Reporting of Observational Studies in Epidemiology (STROBE) and the American Association for Public Opinion Research (AAPOR) guidance. Descriptive statistics are presented as number and percentage (n, %), with percentages calculated based on the relevant subgroup for each analysis. Exploratory Wilcoxon rank-sum test and logistic regression were conducted to examine (1) demographic differences between AI users and non-users, (2) differences in mental health-related chatbot use between participants with and without a current mental health diagnosis, and (3) the relationship between AI use patterns and discussion of chatbot use to mental health providers. Given the descriptive nature of this paper, the comparisons were not conducted to test specific hypotheses and were not adjusted for multiple comparisons. No a priori sample size calculation was conducted. The analysis used all completed responses. Missing data were handled by complete-case analysis. Denominators varied by design based on relevant subgroups (e.g., AI users, mental health users). All analyses were conducted using R version 4.5.2 [7].

## Results

### Participant Characteristics

Out of 523 individuals who accessed the survey, 454 (86.8%) completed the survey (Table 1). Missingness was minimal (< 0.22%). Participant age ranged from 18 to 65 years and older, with approximately half (45.5%) falling between the ages of 30 and 56. The participants were predominantly female (80.1%) and identified primarily as White (78.4%) and non-Hispanic (93.4%). Approximately half of the participants (47.0%) lived in suburban areas and roughly one-third (31.4%) in urban areas, with the remainder in rural settings. Most participants had completed at least some college education (94.0%). With respect to mental health, 64.6% of the participants reported a current mental health diagnosis. Most respondents were affiliated with NAMI (78.0%).

**Table 1.**
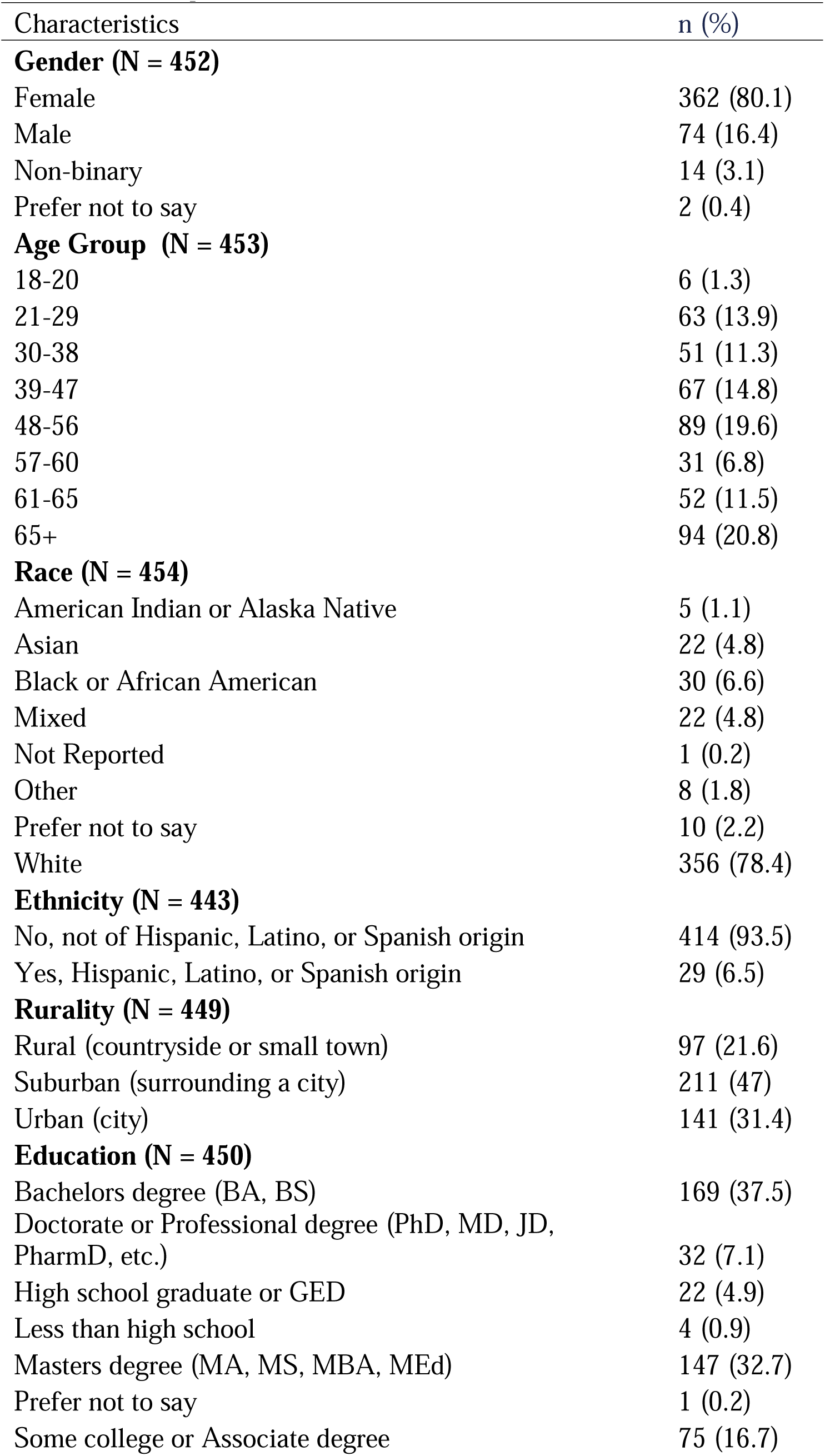
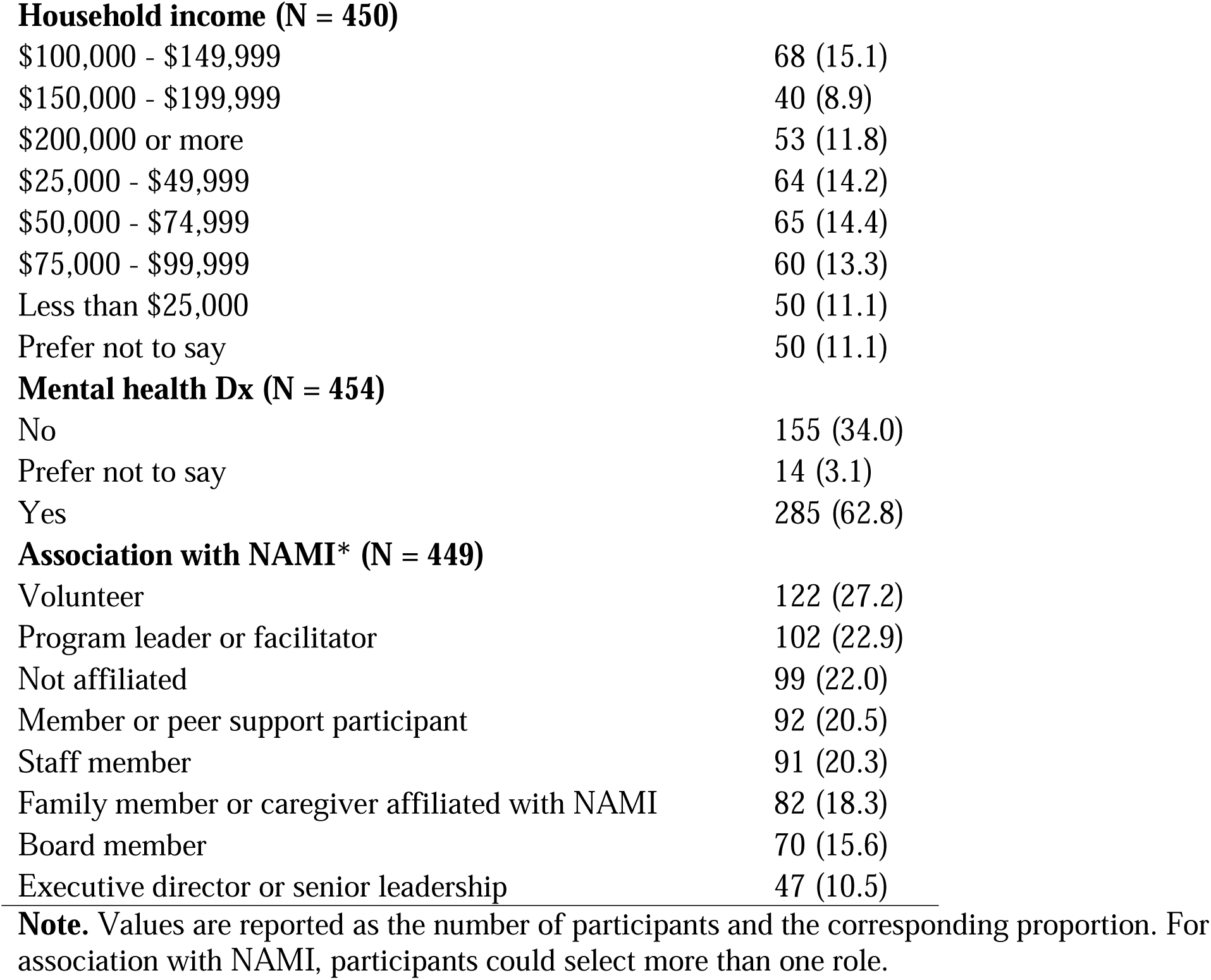
Descriptive statistics.

| Characteristics | n (%) |
| --- | --- |
| **Gender (N = 452)** |  |
| Female | 362 (80.1) |
| Male | 74 (16.4) |
| Non-binary | 14 (3.1) |
| Prefer not to say | 2 (0.4) |
| **Age Group (N = 453)** |  |
| 18-20 | 6 (1.3) |
| 21-29 | 63 (13.9) |
| 30-38 | 51 (11.3) |
| 39-47 | 67 (14.8) |
| 48-56 | 89 (19.6) |
| 57-60 | 31 (6.8) |
| 61-65 | 52 (11.5) |
| 65+ | 94 (20.8) |
| **Race (N = 454)** |  |
| American Indian or Alaska Native | 5 (1.1) |
| Asian | 22 (4.8) |
| Black or African American | 30 (6.6) |
| Mixed | 22 (4.8) |
| Not Reported | 1 (0.2) |
| Other | 8 (1.8) |
| Prefer not to say | 10 (2.2) |
| White | 356 (78.4) |
| **Ethnicity (N = 443)** |  |
| No, not of Hispanic, Latino, or Spanish origin | 414 (93.5) |
| Yes, Hispanic, Latino, or Spanish origin | 29 (6.5) |
| **Rurality (N = 449)** |  |
| Rural (countryside or small town) | 97 (21.6) |
| Suburban (surrounding a city) | 211 (47) |
| Urban (city) | 141 (31.4) |
| **Education (N = 450)** |  |
| Bachelors degree (BA, BS) | 169 (37.5) |
| Doctorate or Professional degree (PhD, MD, JD, PharmD, etc.) | 32 (7.1) |
| High school graduate or GED | 22 (4.9) |
| Less than high school | 4 (0.9) |
| Masters degree (MA, MS, MBA, MEd) | 147 (32.7) |
| Prefer not to say | 1 (0.2) |
| Some college or Associate degree | 75 (16.7) |
| **Household income (N = 450)** |  |
| \$100,000 - \$149,999 | 68 (15.1) |
| \$150,000 - \$199,999 | 40 (8.9) |
| \$200,000 or more | 53 (11.8) |
| \$25,000 - \$49,999 | 64 (14.2) |
| \$50,000 - \$74,999 | 65 (14.4) |
| \$75,000 - \$99,999 | 60 (13.3) |
| Less than \$25,000 | 50 (11.1) |
| Prefer not to say | 50 (11.1) |
| **Mental health Dx (N = 454)** |  |
| No | 155 (34.0) |
| Prefer not to say | 14 (3.1) |
| Yes | 285 (62.8) |
| **Association with NAMI\* (N = 449)** |  |
| Volunteer | 122 (27.2) |
| Program leader or facilitator | 102 (22.9) |
| Not affiliated | 99 (22.0) |
| Member or peer support participant | 92 (20.5) |
| Staff member | 91 (20.3) |
| Family member or caregiver affiliated with NAMI | 82 (18.3) |
| Board member | 70 (15.6) |
| Executive director or senior leadership | 47 (10.5) |

**Note.** Values are reported as the number of participants and the corresponding proportion. For association with NAMI, participants could select more than one role.

Of the 454 participants who completed the survey, 316 (69.6%) reported having ever used an AI chatbot, 113 (24.9%) reported never having used one, and 25 (5.5%) were unsure what the question meant. AI users were significantly younger (median age category: 39-47 years) than non-users (median age category: 48-56 years; *W =* 9617, *P* < .001). Report of AI use was more common among those who reported a current mental health diagnosis than those who did not (77.0% vs 67.3%; OR, 1.60; 95% CI, 1.02-2.49; *P* = .04). There was no significant difference between AI users and non-users in gender, race (White vs non-White), ethnicity, or rurality.

### AI Chatbot Adoption

Among 316 AI users, 196 (62.0%) reported having used more than one chatbot in the past, with an average of 2.10 chatbots per person. ChatGPT was the most widely adopted (Figure 1), with 277 participants (87.7%) having used the tool, followed by Google Gemini at 156 (49.3%). Most AI users (228, 72.2%) did not pay for a chatbot subscription. Among those who did pay for a subscription, ChatGPT was the most commonly subscribed-to tool, reported by 46 participants (14.6%). Over half of participants used AI primarily to support themselves (57.9%), one-third (37.6%) used it for others, and the rest (4.5%) used it for both.

**Figure 1.**
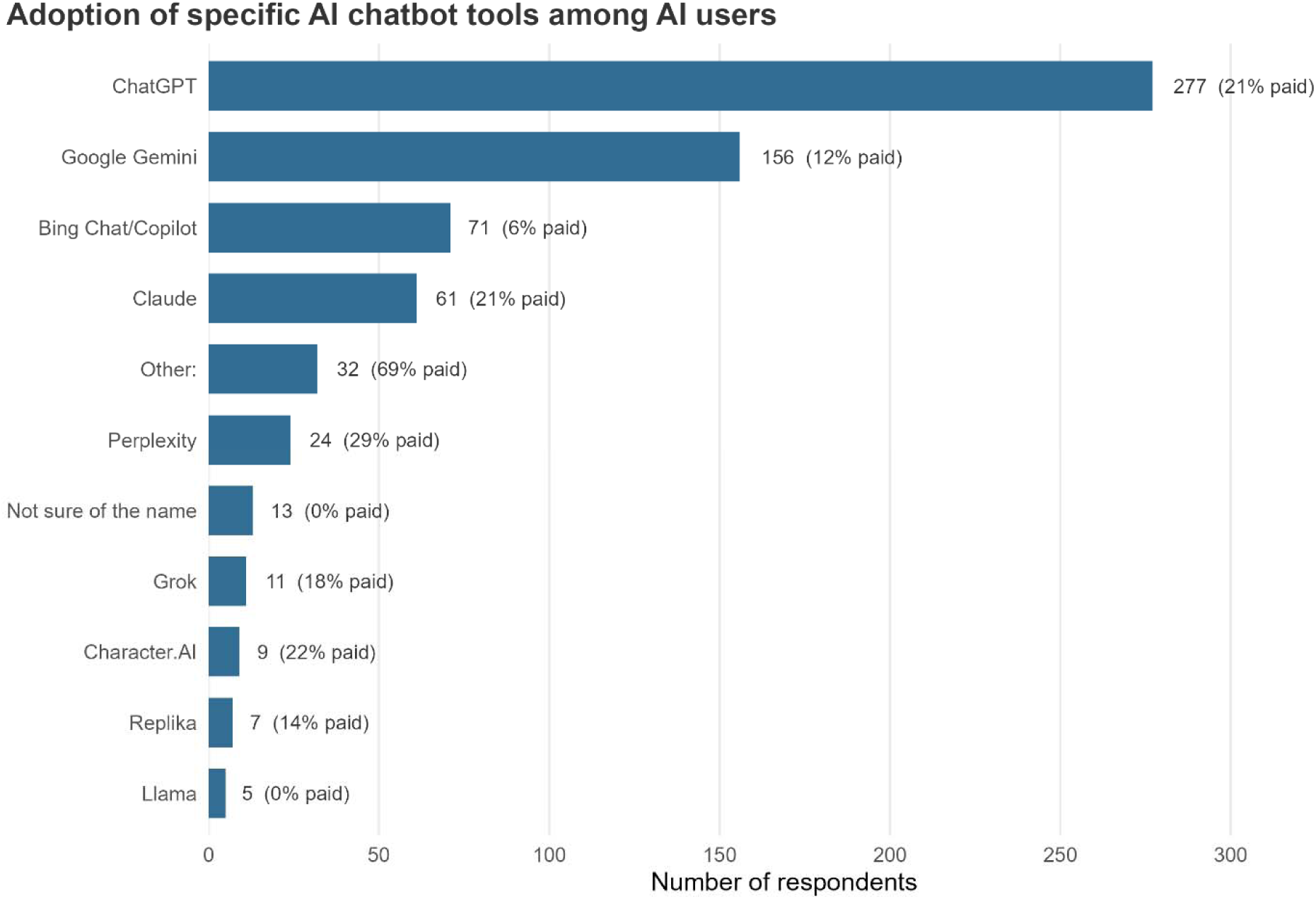
**Note.** N = 316 participants who indicated having used AI tool in the past. Participants were allowed to choose more than one model. The number at the end of each bar reflects the count of respondents who endorsed using that tool and the percentage of those users who reported paying for that tool.

Frequency and duration of chatbot use varied across the participants (Figure 2). Eighty-two participants (25.9%) reported using chatbots daily, 81 (25.6%) several times a week, 52 (16.5%) a few times a month, and 51 (16.1%) rarely. Compared with when the first used AI chatbots, 161 participants (50.9%) indicated using them more frequently, 79 (25.0%) about the same, and 44 (13.9%) less often.

**Figure 2.**
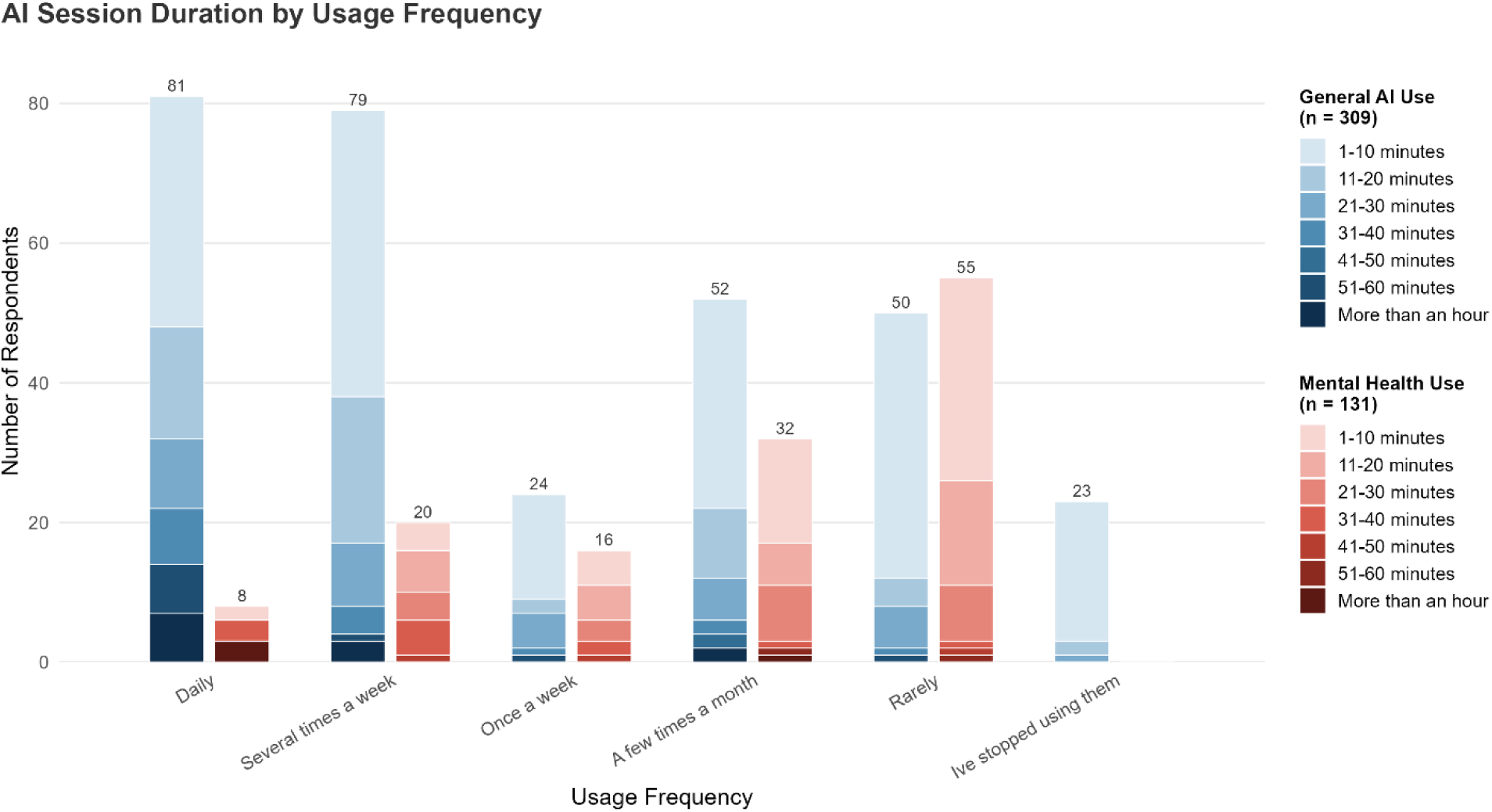
**Note.** Bars are stacked by typical session length. Eight participants who did not report usage frequency or typical session length were excluded from the graph. The option “I’ve stopped using them” was not asked for mental health use.

Session durations also spanned a wide range, with the most commonly reported session length being 1 – 10 minutes (177, 57.1%), 55 participants (17.7%) reported sessions for 11-20 minutes, and 37 (11.9%) for 21-30 minutes. Only 12 (3.9%) reported typical sessions longer than an hour. Half of participants (51.3%) reported starting a new conversation in each session, and 36.1% indicated that it depends on the specific topic.

A total of 305 participants reported their typical purposes for chatbot use. The most commonly reported purpose of chatbot use was fact-finding (similar to internet searching), reported by 220 participants (72.1%), followed by work or administrative tasks (217, 71.1%), and general learning (150, 51.5%).

### Mental Health Related Chatbot Use

Among the 316 AI users, 133 (42.1%) reported having used an AI chatbot for mental health-related purposes. Among those who had not used chatbots for mental health (183, 57.9%), the most commonly cited reasons for not using were a preference for human (cited by 141, 77.0%), mistrust of AI (94, 51.4%), and doubts about effectiveness (91, 49.7%).

Despite a substantial report of AI chatbots for mental health purposes, actual engagement was limited. Only a small proportion reported using AI chatbots for mental health daily (8, 6.1%) or several times a week (20, 15.1%), while a larger proportion reported a few times a month (32, 24.2%,) or rarely (56, 42.4%). Typical session duration was slightly longer for mental health purposes than general use but still brief (Figure 2), with 56 (42.1%) reporting sessions of under 10 minutes, followed by 32 (24.1%) reporting 11 – 20 minutes. In terms of the number of messages exchanged per session, 75 (57.2%) reported sending under 6 messages, 34 (25.9%) reported between 7 and 12 messages.

Participants endorsed a range of specific activities related to mental health (Figure 3). The most commonly endorsed activities were information gathering (cited by 76, 57.1%), managing anxiety or stress (66, 49.6%), and learning about oneself (62, 46.6%). Thirteen participants (9.8%) reported having used an AI chatbot for companionship and 25 (18.8%) reported using one during mental health crisis. Among the latter, 19 (76.0%) found the chatbot helpful in reducing distress, five found it neutral, and one found it made things worse. Eighty participants (60.2%) had not used AI during a mental health crisis, and 25 (18.8%) had never experienced a crisis.

**Figure 3.**
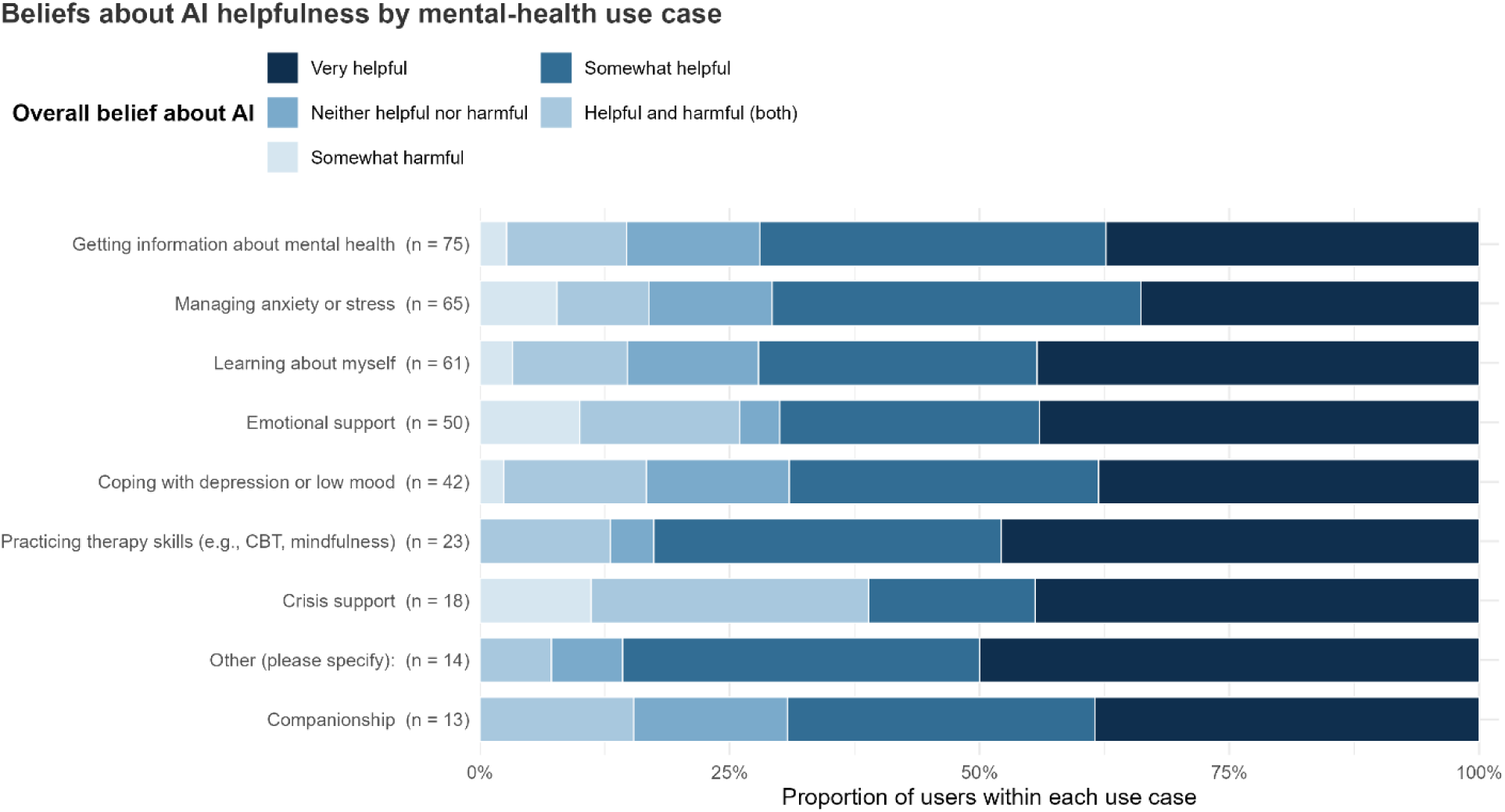
**Note.** Distribution of overall perceived helpfulness of AI chatbots among participants who reported using AI for mental health purposes (N = 130). Three participants who did not report specific use case or helpfulness were excluded. Participants could endorse multiple use cases. Bars are stacked by perceived helpfulness. No participant selected “Very harmful”.

Attitude toward AI use for mental health was neutral to positive. Sixty-one (45.9%) indicated feeling neutral towards the advice, with 49 (36.8%) “mostly trusting”, and 15 (11.3%) “mostly distrusting”. Most participants (91, 68.4%) rated the advice to be comparable to an internet search or a friend’s advice. Fifty-eight (43.6%) would recommend AI chatbots “with caution” to others with mental health condition, 30 (22.6%) “probably not” and 21 (15.8%) were “unsure”. Only 14 (10.5%) indicated that they would “definitely” recommend AI chatbots to others with mental health conditions.

Many perceived AI chatbots as helpful for mental health: 43 (32.3%) rated them as “very helpful”, 47 (35.3%) as “somewhat helpful”, and 16 (12.0%) as both helpful and harmful (Figure 3). Only 6 (4.5%) rated “somewhat harmful”, and no participant selected “very harmful”. The 125 participants who rated the tools as neutral or helpful were asked to identify specific ways AI chatbots were helpful. Most of the provided options were chosen by roughly half of these participants, with “available 24/7” (endorsed by 83, 66.4%) and “cost-effective” (75, 60.0%) being the most frequently endorsed. Among 41 participants who rated the tools as neutral or harmful, specific reasons for the perceived harm included “didn’t understand my situation” (endorsed by 18, 43.9%), “became a way to avoid professional help” (14, 34.1%).

Discussion with mental health providers about AI use was limited. Among the 95 participants with a mental health provider, only 14 (14.7%) had discussed their AI use openly with their provider, and 25 (26.3%) had mentioned it briefly. A larger proportion (56, 58.9%) indicated not having had the discussion, and among them, 43 indicated not planning to have a discussion. Higher frequency of AI use was associated with greater odds of disclosure to providers (OR per level of increase in frequency, 1.67; 95% CI, 1.20–2.38; *P* = .003). Length of AI sessions and number of messages sent per session were not significantly associated with disclosure of chatbot use.

Participants who reported a current mental health diagnosis were more likely to report using an AI chatbot for mental health than those without a diagnosis (46.1% vs 32.7%; OR 1.76; 95% CI, 1.04 -3.00; *P* = .03). However, diagnosis status was not associated with frequency or duration of use, use during a crisis, trust in chatbots, perceived helpfulness, or disclosure of chatbot use to mental health providers.

### Perspectives of Non-Users

Among the 113 participants who reported never having used an AI chatbot for any purpose, the most commonly cited reasons for non-use were preference for human interaction (cited by 67, 59.3%), mistrust of AI (52, 46.0%), lack of need (52, 46.0). A separate question asked why they would not use a chatbot for mental health purpose specifically (Figure 4). The most commonly endorsed reasons were preference for human interaction (cited by 77, 68.1%), mistrust of AI with sensitive information (52, 46.0%), privacy/security concern (48, 42.5%).

**Figure 4.**
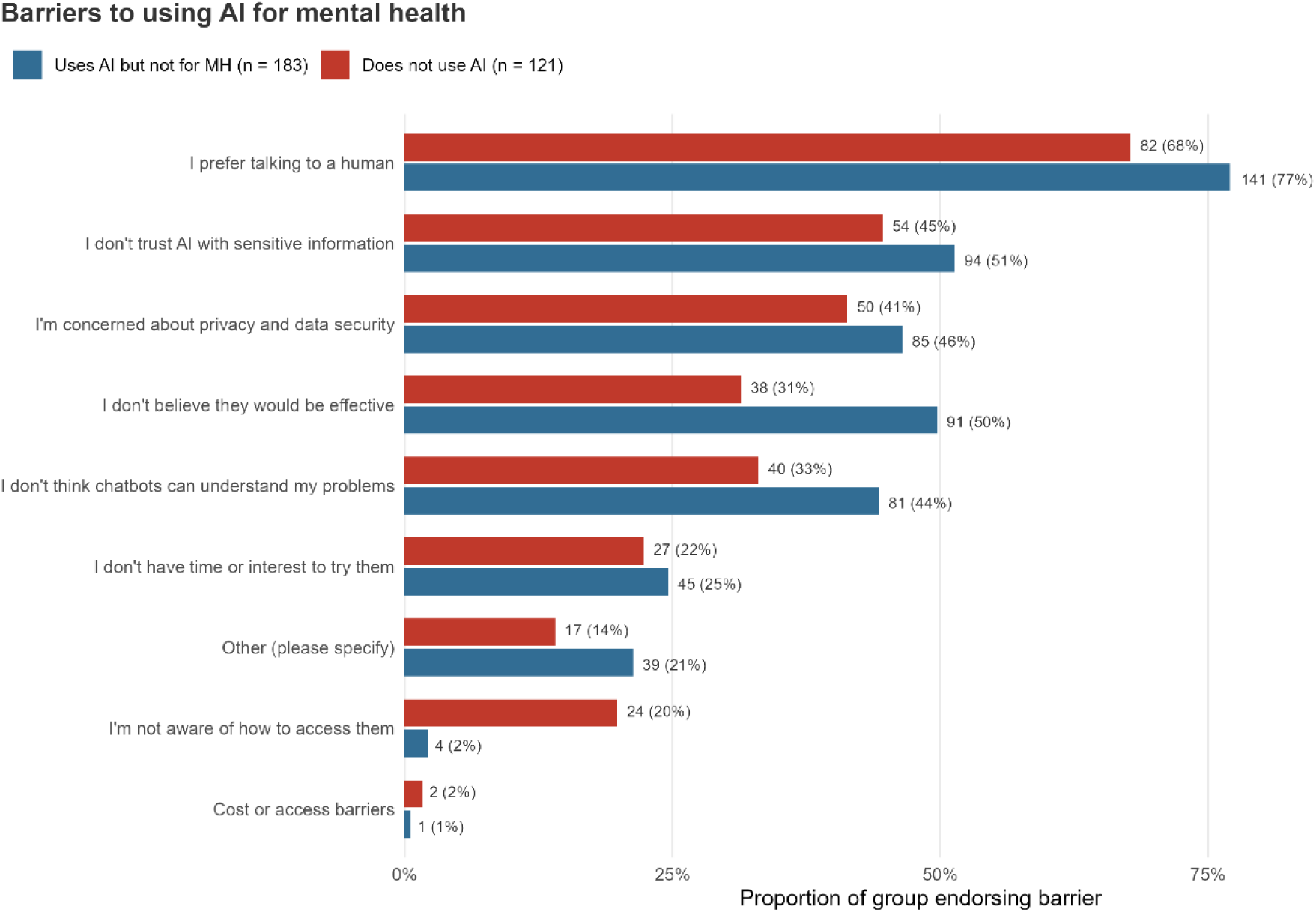
**Note.** Self-reported barriers to using AI for mental health, compared between respondents who uses AI but not for mental health, versus those who do not use AI at all. Percentages reflect the proportion of each group endorsing each barrier. Participants could select multiple barriers.

When asked about what they considered the best use of AI for mental health, the most commonly endorsed were information gathering (64, 56.6%), practicing therapy skills (28, 24.8%), and managing anxiety or stress (16, 14.1%). Interestingly, the top two endorsed applications (information gathering and anxiety management) paralleled the most common actual uses reported by mental health users. However, the third most common actual use (learning about oneself) was the least endorsed potential use among non-users (6, 5.3%). Finally, 45 of non-users (40.2%) reported interest in learning more about AI chatbots, suggesting that a meaningful proportion of current non-users are open to future engagement.

## Discussion

The present study explored AI chatbot adoption and mental health-related use among adults connected to a large nonprofit mental health organization in the US Three findings emerged from the survey. First, although AI chatbots were used for mental health purposes, the pattern of use was heterogeneous. Many users reported only infrequent use, with purposes centering on information seeking and in-the-moment coping rather than deeper or more sustained forms of connection or therapy. Second, users generally trusted AI chatbots and described them as predominantly helpful for mental health. Third, although AI use for mental health was more common among participants with a mental health diagnosis, usage patterns or perceived helpfulness did not differ meaningfully from those without a diagnosis. These findings paint a nuanced picture of AI use for mental health.

We observed a higher prevalence of AI use for mental health (42.1%) than reported in general population poll (16%) [1], but was comparable to a study that explicitly targeted adults with mental health diagnosis (48.7%) [8]. This pattern is consistent with our finding that participants with a diagnosis were more likely to use AI chatbot for mental health than those without. Beyond prevalence, our results extend prior work by characterizing the intensity and nature of AI use for mental health. Most users reported having short, few-turn conversations focused on information seeking and in-the-moment coping, and that they find these interactions helpful. On the other hand, potentially concerning use cases such as companionship or crisis support and experiences of harm were rare although present. The finding highlights an important limitation in previous reporting: pooled prevalence may overestimate how the majority actually use AI chatbots, while underestimating the intense use among a smaller group of heavy users. Mirroring the prevalence of AI tool use, most participants reported using general-purpose chatbots such as ChatGPT rather than mental-health-specific chatbots. While recent work suggests that general-purpose chatbots may perform comparably to mental health chatbots [10], the effectiveness of both general-purpose and mental health-specific chatbots remains underexplored. Similarly, a recent systematic review and meta-analysis suggested that self-guided AI chatbots for mental health may have the same effect size as mental health apps [11], and raises the need for a new generation of higher-quality studies. Given the reported high rates of general-purpose chatbots for mental health, the need for head-to-head studies of mental health vs general-purpose AI chatbots across both safety and effectiveness, as well as digital placebo-controlled studies, is urgent.

Finally, our results suggested that discussion with mental health providers about AI usage was rare. Clinicians may therefore be unaware of an important source of mental health input or coping strategies that may conflict with clinical recommendations [12]. Future work should examine barriers to disclosure and identify ways that clinicians may proactively ask patients about AI use in routine care, and the patients are comfortable sharing use.

The study’s partnership with NAMI allowed us to reach adults with experience with mental health, addressing a methodological gap in the literature that frequently relied on crowdsourced samples. However, several limitations should be noted. First, all measures, including reports of diagnoses, are self-reported. Second, the cross-sectional design precludes inference about change over time or causality. Third, comparisons were exploratory with no a priori hypothesis or correction for multiple comparisons and should be interpreted descriptively. Fourth, the survey was distributed across multiple overlapping NAMI mailing lists. Because the number of unique recipients could not be determined, we were unable to calculate the response rate. To address this limitation, we characterized the demographic composition of the participants in detail. The distribution of the demographic composition was consistent with prior studies of NAMI recruited participants [13,14], suggesting that our sample is likely representative of the broader NAMI community. Nevertheless, while NAMI is one of the largest mental health organizations, participants were from a single organization and is not representative of the broader population. The use of AI for mental health, by people with actual mental health needs, continues to expand. However, today this use remains heterogeneous, brief, and largely invisible to clinicians, suggesting that AI is functioning less as a replacement for therapy than as an accessible, around-the-clock tool for information seeking and immediate coping. As regulators, clinicians, and technology vendors seek to make these tools safer, generating rigorous, real-world evidence from actual patient populations, rather than crowdsourced proxies, is the necessary foundation.

## Data Availability

Individual participant data are not available per the terms of participant consent.

## Author Contributions

Concept and design: JT, LW, CC, MW, DG

Acquisition, analysis, interpretation of data: HN, SR, JT, PN, MF, JN

Drafting of the manuscript: HN, JT

Critical review of the manuscript: LW, CC, MW, DG, SR, PN, MF, JN, JT

Final approval of the manuscript: all authors

JT had full access to all of the data in the study and takes responsibility for the integrity of the work as a whole. All authors agree to be accountable for all aspects of the work and to ensure that questions related to the accuracy or integrity of the work are appropriately investigated and resolved.

## Data Sharing Statement

Individual participant data are not available per the terms of participant consent.

During the preparation of this work, the authors used Claude Opus 4.8 to assist with editing sentences for grammar, clarity, and flow. The tool was not used to generate ideas, conduct a literature review, design or carry out research, analyze data, or write any section of the manuscript independently. After using this tool, the authors reviewed and edited the content and take full responsibility for the content of the published article.

## Reference

1. Montero A, Montalvo J III, Kearney A, Valdes I, Kirzinger A, Hamel L. KFF Tracking Poll on Health Information and Trust: Use of AI for Health Information and Advice. KFF. Published March 25, 2026. Accessed June 29, 2026. https://www.kff.org/public-opinion/kff-tracking-poll-on-health-information-and-trust-use-of-ai-for-health-information-and-advice/

2. McClain C, Anderson M, Sidoti O, Bishop W. How teens use and view AI. Pew Research Center. Published February 24, 2026. Accessed June 29, 2026. https://www.pewresearch.org/internet/2026/02/24/how-teens-use-and-view-ai/

3. Inkster B, Sarda S, Subramanian V. An empathy-driven, conversational artificial intelligence agent (Wysa) for digital mental well-being: real-world data evaluation mixed-methods study. JMIR Mhealth Uhealth. 2018;6(11):e12106. doi:10.2196/12106

4. Rousmaniere T, Zhang Y, Li X, Shah S. Large language models as mental health resources: patterns of use in the United States. Pract Innov. 2026;11(2):139–155. doi:10.1037/pri0000292

5. Stade EC, Stirman SW, Ungar LH, et al. Large language models could change the future of behavioral healthcare: a proposal for responsible development and evaluation. Npj Ment Health Res. 2024;3(1):12. doi:10.1038/s44184-024-00056-z

6. Lisowski V, Shumate JN, Yassin W, Song SH, Kalinich M, Ram N, Lee A, Flathers M, Ryan S, Keshavan M, Torous J. A self-reported survey on the use of AI chatbots by people on an inpatient psychiatric unit. Psychiatric Services. Under review

7. R Core Team. R: A Language and Environment for Statistical Computing. Version 4.x. R Foundation for Statistical Computing; 2026. Accessed June 29, 2026. https://www.R-project.org/

8. Rousmaniere T, Zhang Y, Li X, Shah S. Large language models as mental health resources: patterns of use in the United States. Pract Innov. Published online 2025. doi:10.1037/pri0000292

9. Coombs NC, Meriwether WE, Caringi J, Newcomer SR. Barriers to healthcare access among US adults with mental health challenges: a population-based study. SSM Popul Health. 2021;15:100847. doi:10.1016/j.ssmph.2021.100847

10. Kuta B, Novak L, Zidkova R, et al. Effectiveness of a fully automated mobile therapeutic versus a general chatbot in reducing depression and anxiety and improving well-being: feasibility randomized controlled trial. JMIR Ment Health. 2026;13:e82642. doi:10.2196/82642

11. Yang H, Chang F, Muroi F, Liu Z, Zhang W, Cai J. Commercial AI-based mental health chatbots as low-intensity adjuncts to psychotherapy: effectiveness, adherence, and safety—a systematic review and meta-analysis. Psychother Psychosom. Published online 2026

12. Saba SK, Weeks WB. Patients use AI—clinicians should ask how. JAMA Psychiatry. 2026;83(5):543–544. doi:10.1001/jamapsychiatry.2026.0451

13. Marshall TB, Solomon P. Releasing information to families of persons with severe mental illness: a survey of NAMI members. Psychiatr Serv. 2000;51(8):1006–1011. doi:10.1176/appi.ps.51.8.1006

14. Smith ME, Lindsey MA, Williams CD, et al. Race-related differences in the experiences of family members of persons with mental illness participating in the NAMI Family to Family Education Program. Am J Community Psychol. 2014;54(3-4):316–327.

