## Appendix 1 for "Use and Perceptions of AI Chatbots for Mental Health Support Among Adults with Lived Experience"

#### Demographics

| | |
|---|---|
| Please select your age range | ◯ 18-20<br>◯ 21-29<br>◯ 30-38<br>◯ 39-47<br>◯ 48-56<br>◯ 57-60<br>◯ 61-65<br>◯ 65+ |
| What is your Gender Identity? | ◯ Female<br>◯ Male<br>◯ Non-binary<br>◯ Prefer to self-describe<br>◯ Prefer not to say |
| Please describe: | __________________________________ |
| Are you of Hispanic, Latino, or Spanish origin? | ◯ No, not of Hispanic, Latino, or Spanish origin<br>◯ Yes, Hispanic, Latino, or Spanish origin |
| What is your race? (Select all that apply) | ☐ American Indian or Alaska Native<br>☐ Asian<br>☐ Black or African American<br>☐ Middle Eastern or North African<br>☐ Native Hawaiian or Other Pacific Islander<br>☐ White<br>☐ Other race: _____<br>☐ Prefer not to say |
| Please enter your race: | __________________________________ |
| How would you describe the area where you currently live? | ◯ Urban (city)<br>◯ Suburban (surrounding a city)<br>◯ Rural (countryside or small town) |
| What is your highest level of education? | ◯ Less than high school<br>◯ High school graduate or GED<br>◯ Some college or Associate degree<br>◯ Bachelor's degree (BA, BS)<br>◯ Master's degree (MA, MS, MBA, MEd)<br>◯ Doctorate or Professional degree (PhD, MD, JD, PharmD, etc.)<br>◯ Prefer not to say |
| What is your current annual household income? | ◯ Less than $25,000<br>◯ $25,000 - $49,999<br>◯ $50,000 - $74,999<br>◯ $75,000 - $99,999<br>◯ $100,000 - $149,999<br>◯ $150,000 - $199,999<br>◯ $200,000 or more<br>◯ Prefer not to say |

| | |
|---|---|
| What is your relationship to the National Alliance on Mental Illness (NAMI)? Select all that apply | ☐ Executive Director or senior leadership<br>☐ Staff member<br>☐ Program leader or facilitator<br>☐ Board member<br>☐ Volunteer<br>☐ Member or peer support participant<br>☐ Family member or caregiver affiliated with NAMI<br>☐ I am not affiliated with NAMI |
| Do you currently have a mental health diagnosis? | ◯ Yes<br>◯ No<br>◯ Prefer not to say |
| Have you ever used an AI chatbot or language model? | ◯ Yes<br>◯ No<br>◯ Not sure what this means |
| We use the term 'AI chatbot' to refer to tools like ChatGPT, Gemini, Claude, Grok, or Copilot. These are websites, apps, or software you can have a text conversation with to get information, help, or support. With that in mind, have you used any of these tools? | ◯ Yes, I have used one of these tools<br>◯ No, I have not used any of these tools<br>◯ I'm still not sure |

Progress: 15%

### AI Users

AI Use

| Question | Options |
| --- | --- |
| Which AI chatbot tools have you used? (Check all that apply) | ☐ ChatGPT<br>☐ Perplexity<br>☐ Claude<br>☐ Google Gemini<br>☐ Grok<br>☐ Bing Chat/Copilot<br>☐ Llama<br>☐ Character.AI<br>☐ Replika<br>☐ Not sure of the name<br>☐ Other: |
| Which other AI chatbot tools have you used? | __________________________________ |
| Which AI chatbot tools do you pay for? (Check all that apply) | ☐ ChatGPT<br>☐ Perplexity<br>☐ Claude<br>☐ Google Gemini<br>☐ Grok<br>☐ Bing Chat/Copilot<br>☐ Llama<br>☐ Character.AI<br>☐ Replika<br>☐ Other:<br>☐ I don't pay for any |
| Which other AI chatbot tool do you pay for? | __________________________________ |
| How often do you currently use AI chatbots? | ◯ Daily<br>◯ Several times a week<br>◯ Once a week<br>◯ A few times a month<br>◯ Rarely<br>◯ I've stopped using them |
| When you use AI, how long is a typical session? | ◯ 1-10 minutes<br>◯ 11-20 minutes<br>◯ 21-30 minutes<br>◯ 31-40 minutes<br>◯ 41-50 minutes<br>◯ 51-60 minutes<br>◯ More than an hour |
| When you use AI chatbots, do you typically start a new chat or continue a previous chat thread? | ☐ I usually start a new chat each time<br>☐ I usually continue a previous chat thread<br>☐ It depends<br>☐ I'm unsure |
| Compared to when you first tried AI chatbots, how has your usage changed? | ◯ I use them more often now<br>◯ I use them about the same amount<br>◯ I use them less often now<br>◯ I stopped using them |

If you've stopped using AI chatbots, please answer the remaining questions based on your past experience.

| | |
|---|---|
| What do you typically use AI chatbots for? Select all that apply. | ☐ Mental Health<br>☐ Emotional support (venting, ranting, symptom management)<br>☐ Emotional processing (processing of emotions and feelings)<br>☐ Identifying my emotions<br>☐ Clinical information/advice (diagnosis /psychoeducation)<br>☐ Therapeutic exercises (CBT practice, exposure simulation)<br>☐ Crisis support<br>☐ Wellness (e.g. exercise, meal prepping, yoga, spirituality)<br>☐ Communication skills and practice<br>☐ Romantic/Sexual communication<br>☐ Fact finding and internet searching<br>☐ Learning<br>☐ Digital small talk (general topic conversation)<br>☐ Work/administrative support and tasks<br>☐ Content generation (images, art, writing, code) |
| Beyond what's listed above, what else do you use AI chatbots for? | ______________________________ |
| When you use AI chatbots, are you typically seeking information or support for yourself or others? | ○ Myself<br>○ Other people<br>○ Both |

Progress: 40%

#### Mental Health Related Use

Have you ever used an AI chatbot for your mental health?

By mental health, we mean any way you use AI to change how you feel, think, or connect. These may include:

- Emotional support (venting, expressing frustration, symptom management)
- Emotional processing or self-reflection (understanding and working through your feelings)
- Clinical information/advice (diagnosis or education about symptoms)
- Practicing coping skills or therapy homework
- Companionship
- Getting information about mental health
- Crisis support

◯ Yes (frequently)
◯ Yes (occasionally)
◯ Yes (once or twice)
◯ No/never

How have you used AI chatbots for your mental health? (Check all that apply)

☐ Emotional support
☐ Companionship
☐ Managing anxiety or stress
☐ Coping with depression or low mood
☐ Getting information about mental health
☐ Practicing therapy skills (e.g., CBT, mindfulness)
☐ Learning about myself
☐ Crisis support
☐ Other (please specify):

Please specify: ______________________________

Which of the following describe why you have not used AI chatbots for mental health? (Check all that apply)

☐ I don't trust AI with sensitive information
☐ I'm concerned about privacy and data security
☐ I prefer talking to a human
☐ I don't think chatbots can understand my problems
☐ I'm not aware of how to access them
☐ I don't believe they would be effective
☐ I don't have time or interest to try them
☐ Cost or access barriers
☐ Other (please specify):

Please specify: ______________________________

When you use AI chatbots for mental health, how long do you typically spend in a single session?

◯ 1-10 minutes
◯ 11-20 minutes
◯ 21-30 minutes
◯ 31-40 minutes
◯ 41-50 minutes
◯ 51-60 minutes
◯ More than an hour
(A session means one sitting from when you open the chatbot to when you stop using it.)

| | |
|---|---|
| How many messages do you typically send in a single session? (Count only your own messages, not the AI's responses) | ◯ 1-3<br>◯ 4-6<br>◯ 7-9<br>◯ 10-12<br>◯ 13-15<br>◯ 16-18<br>◯ 19-21<br>◯ 22-24<br>◯ 25-27<br>◯ 28-30<br>◯ 31+ |
| How often do you use AI chatbots specifically for mental health purposes? | ◯ Daily<br>◯ Several times a week<br>◯ Once a week<br>◯ A few times a month<br>◯ Rarely |
| Overall, do you believe that using AI chatbots has been helpful or harmful to your mental health? | ◯ Very helpful<br>◯ Somewhat helpful<br>◯ Neither helpful nor harmful<br>◯ Helpful and harmful (both)<br>◯ Somewhat harmful<br>◯ Very harmful |
| In what ways have AI chatbots been HELPFUL? (Check all that apply) | ☐ Available 24/7 when I need support<br>☐ Cost-effective<br>☐ Non-judgmental space to express feelings<br>☐ Helped me understand my symptoms better<br>☐ Provided coping strategies<br>☐ Helped with cognitive flexibility and insight<br>☐ Helped me prepare for therapy sessions<br>☐ Reduced feelings of isolation<br>☐ Help me prepare for a real-life situation (e.g., role-play, scripting conversations)<br>☐ Other: |
| If other, please explain: | __________________________________ |
| In what ways have AI chatbots been HARMFUL? (Check all that apply) | ☐ Gave incorrect or concerning advice<br>☐ Gave correct but dangerous/unsafe advice<br>☐ Made me feel more isolated from real people<br>☐ Increased my symptoms or distress<br>☐ Became a way to avoid professional help<br>☐ Created dependency or overuse<br>☐ Didn't understand my situation<br>☐ Invalidated my experience<br>☐ Other: |
| If other, please explain. | __________________________________ |
| Have you ever used an AI chatbot when you were in crisis?<br><br>A mental health crisis is when you are experiencing so much distress that you are concerned about your own safety, such as having thoughts of killing yourself, harming yourself, or not being able to take care of your basic needs. | ◯ Yes<br>◯ No<br>◯ Prefer not to say<br>◯ Have not experienced a crisis |

| | |
|---|---|
| If yes, how did the experience affect your crisis? | ◯ Helped reduce distress<br>◯ No effect<br>◯ Made things worse |
| How much do you trust the mental health advice provided by AI chatbots? | ◯ I strongly trust the advice<br>◯ I mostly trust the advice<br>◯ I feel neutral towards the advice<br>◯ I mostly distrust the advice<br>◯ I strongly distrust the advice |
| How does your level of trust in AI chatbots compare to your trust in other sources of mental health information?<br>My trust in mental health information from an AI chatbot is most similar to the level of trust I have for ... | ◯ A licensed mental health provider or doctor<br>◯ An established mental health organization (e.g., NAMI, SAMHSA)<br>◯ A friend<br>◯ An internet search<br>◯ A social media post |

Progress: 60%

#### Relationship with Treatment

| | |
|---|---|
| Have you discussed your AI chatbot use with your mental health providers? | ◯ Yes, I've discussed it openly<br>◯ I've mentioned it briefly<br>◯ No, but I plan to<br>◯ No, and I don't plan to<br>◯ I do not have a mental health provider |
| Would you recommend AI chatbots to other people dealing with mental health challenges? | ◯ Yes, definitely<br>◯ Yes, with caution<br>◯ Unsure<br>◯ Probably not<br>◯ Definitely not |
| What do you think the best uses (if any) for an AI chatbot would be: | ☐ Emotional support<br>☐ Companionship<br>☐ Managing anxiety or stress<br>☐ Coping with depression or low mood<br>☐ Getting information about mental health<br>☐ Practicing therapy skills (e.g., CBT, mindfulness)<br>☐ Knowing about myself<br>☐ Crisis support<br>☐ Other: |
| Please describe: | ______________________________ |

Progress: 75%

### Non Users

#### Non-Users

| | |
|---|---|
| Which of the following describe why you have not used AI chatbots? (Select all that apply) | ☐ Don't know how to access them<br>☐ Don't have the technology/internet<br>☐ Privacy/data concerns<br>☐ Don't trust AI<br>☐ My healthcare team discouraged it<br>☐ Other people discouraged it<br>☐ I am worried it might make my symptoms worse<br>☐ Prefer human interaction<br>☐ Never felt the need<br>☐ Didn't know they existed<br>☐ Cost or access barriers<br>☐ Other (please specify): ________ |
| Please describe: | __________________________________ |
| Which of the following describe why you have not used AI chatbots for mental health? (Select all that apply) | ☐ I don't trust AI with sensitive information<br>☐ I'm concerned about privacy and data security<br>☐ I prefer talking to a human<br>☐ I don't think chatbots can understand my problems<br>☐ I'm not aware of how to access them<br>☐ I don't believe they would be effective<br>☐ I don't have time or interest to try them<br>☐ Cost or access barriers<br>☐ Other (please specify): ________ |
| Please describe: | __________________________________ |
| What do you think the best uses (if any) for an AI chatbot would be? (Check all that apply) | ☐ Emotional support<br>☐ Companionship<br>☐ Managing anxiety or stress<br>☐ Coping with depression or low mood<br>☐ Getting information about mental health<br>☐ Practicing therapy skills (e.g., CBT, mindfulness)<br>☐ Knowing about myself<br>☐ Crisis support |
| Would you be interested in learning more about AI chatbots? | ◯ Yes<br>◯ No |

Progress: 75%

STROBE Statement—Checklist of items that should be included in reports of ***cross-sectional studies***

| | Item No | Recommendation | Page No |
|---|---|---|---|
| **Title and abstract** | 1 | (*a*) Indicate the study's design with a commonly used term in the title or the abstract | 2 |
| | | (*b*) Provide in the abstract an informative and balanced summary of what was done and what was found | 2 |
| **Introduction** | | | |
| Background/rationale | 2 | Explain the scientific background and rationale for the investigation being reported | 4 |
| Objectives | 3 | State specific objectives, including any prespecified hypotheses | 5 |
| **Methods** | | | |
| Study design | 4 | Present key elements of study design early in the paper | 5 |
| Setting | 5 | Describe the setting, locations, and relevant dates, including periods of recruitment, exposure, follow-up, and data collection | 6 |
| Participants | 6 | (*a*) Give the eligibility criteria, and the sources and methods of selection of participants | 5 |
| Variables | 7 | Clearly define all outcomes, exposures, predictors, potential confounders, and effect modifiers. Give diagnostic criteria, if applicable | NA |
| Data sources/ measurement | 8* | For each variable of interest, give sources of data and details of methods of assessment (measurement). Describe comparability of assessment methods if there is more than one group | *6* |
| Bias | 9 | Describe any efforts to address potential sources of bias | NA |
| Study size | 10 | Explain how the study size was arrived at | NA |
| Quantitative variables | 11 | Explain how quantitative variables were handled in the analyses. If applicable, describe which groupings were chosen and why | 6 |
| Statistical methods | 12 | (*a*) Describe all statistical methods, including those used to control for confounding | 6 |
| | | (*b*) Describe any methods used to examine subgroups and interactions | 6 |
| | | (*c*) Explain how missing data were addressed | 6 |
| | | (*d*) If applicable, describe analytical methods taking account of sampling strategy | NA |
| | | (*e*) Describe any sensitivity analyses | NA |
| **Results** | | | |
| Participants | 13* | (a) Report numbers of individuals at each stage of study—eg numbers potentially eligible, examined for eligibility, confirmed eligible, included in the study, completing follow-up, and analysed | 7 |
| | | (b) Give reasons for non-participation at each stage | NA |
| | | (c) Consider use of a flow diagram | NA |
| Descriptive data | 14* | (a) Give characteristics of study participants (eg demographic, clinical, social) and information on exposures and potential confounders | 7 |
| | | (b) Indicate number of participants with missing data for each variable of interest | 7 |
| Outcome data | 15* | Report numbers of outcome events or summary measures | NA |
| Main results | 16 | (*a*) Give unadjusted estimates and, if applicable, confounder-adjusted estimates and their precision (eg, 95% confidence interval). Make clear which confounders were adjusted for and why they were included | 7-11 |

| | | | |
|---|---|---|---|
| | | (*b*) Report category boundaries when continuous variables were categorized | 7-11 |
| | | (*c*) If relevant, consider translating estimates of relative risk into absolute risk for a meaningful time period | NA |
| Other analyses | 17 | Report other analyses done—eg analyses of subgroups and interactions, and sensitivity analyses | 7-11 |
| **Discussion** | | | |
| Key results | 18 | Summarise key results with reference to study objectives | 11 |
| Limitations | 19 | Discuss limitations of the study, taking into account sources of potential bias or imprecision. Discuss both direction and magnitude of any potential bias | 13 |
| Interpretation | 20 | Give a cautious overall interpretation of results considering objectives, limitations, multiplicity of analyses, results from similar studies, and other relevant evidence | 12 |
| Generalisability | 21 | Discuss the generalisability (external validity) of the study results | |
| **Other information** | | | |
| Funding | 22 | Give the source of funding and the role of the funders for the present study and, if applicable, for the original study on which the present article is based | 14 |

*Give information separately for exposed and unexposed groups.

**Note:** An Explanation and Elaboration article discusses each checklist item and gives methodological background and published examples of transparent reporting. The STROBE checklist is best used in conjunction with this article (freely available on the Web sites of PLoS Medicine at http://www.plosmedicine.org/, Annals of Internal Medicine at http://www.annals.org/, and Epidemiology at http://www.epidem.com/). Information on the STROBE Initiative is available at www.strobe-statement.org.
